# Allele frequency differences of causal variants have a major impact on low cross-ancestry portability of PRS

**DOI:** 10.1101/2022.10.21.22281371

**Authors:** Marie Saitou, Andy Dahl, Qingbo Wang, Xuanyao Liu

**Author notes:** The authors contributed equally (Marie Saitou, Andy Dahl).

## Abstract

Genome-wide association studies (GWAS) are overwhelmingly biased toward European ancestries. Nearly all existing studies agree that transferring genetic predictions from European ancestries to other populations results in a substantial loss of accuracy. This is commonly referred to as low portability of polygenic risk scores (PRS) and is one of the most important barriers to the ethical clinical deployment of PRS. Yet, it remains unclear how much various genetic factors, such as linkage disequilibrium (LD) differences, allele frequency differences or causal effect differences, contribute to low PRS portability. In this study, we used gene expression levels in lymphoblastoid cell lines (LCLs) as a simplified model of complex traits with minimal environmental variation, in order to understand how much each genetic factor contributes to PRS portability from European to African populations. We found that *cis-*genetic effects on gene expression are highly similar between European and African individuals (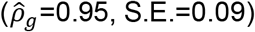). This stands in stark contrast to the very low estimates of *cis*-genetic correlation between Europeans and Africans in previous studies, which we demonstrate are artifacts of statistical bias. We showed that portability decreases with increasing LD differences in the *cis*-regions. We also found that allele frequency differences of causal variants have a striking impact on PRS portability. For example, PRS portability is reduced by more than 32% when the causal *cis*-variant is common (minor allele frequency, MAF > 5%) in European samples (training population) but is rarer (MAF < 5%) in African samples (prediction population). While large allele frequency differences can decrease PRS portability through increasing LD differences, we also show that causal allele frequency can significantly impact portability independently of LD. This observation suggests that improving statistical fine-mapping alone does not overcome the loss of portability caused by causal allele frequency differences. Lastly, we also found that causal allele frequency is the main genetic factor underlying differential gene expression levels across ancestries. We conclude that causal genetic effects are highly similar in Europeans and Africans, and low PRS portability is primarily due to allele frequency differences.

## Background

Many common diseases have genetic basis. Genome-wide association studies (GWAS) were designed to identify genetic risk alleles of common diseases by performing population-based genetic profiling, which has identified thousands of genetic loci associated with complex traits and diseases^1–3^. Disease-associated loci and their effects can be summed to generate polygenic risk scores (PRS), which are used to predict complex traits and disease risk in genotyped individuals. Genetic risk prediction has the potential to enable earlier and tailored disease prevention, particularly if the ancestry of the genotyped individual resembles the ancestry of the GWAS samples. However, the majority of GWAS participants are of European ancestry, which leads to poor prediction accuracy of complex diseases in non-European populations^4,5^. This is known as low cross-ancestry PRS portability. Improving cross-ancestry PRS portability is essential to equitable clinical use of PRS.

The limited portability of cross-ancestry PRS can be attributed to several genetic differences across ancestries, such as linkage disequilibrium (LD) differences, causal allele frequency differences or genetic effect differences. Differential LD between causal genetic variants and the variants used in building PRS (PRS variants) is among one of most well-studied factors lowering portability. Several studies over the past few years have focused on improving PRS portability by developing better statistical methods to overcome the LD differences and more accurately fine-mapping causal genetic variants^6,7^. One recent study^8^ estimated that differences in LD and minor allele frequencies (MAF) of PRS variants together could explained 70-80% of loss of portability from European GWAS to African samples. However, it remains unknown whether and to what extent differences in causal genetic effects and causal allele frequency contribute to low portability. Knowing the contribution of these factors is important because it will inform sampling strategies for future GWAS and aid in development of new statistical methods. For example, if LD contributes to low portability, improved statistical fine-mapping should improve portability. Whereas, if allele frequency or effect size differences are found to affect portability independent of LD, other strategies, such as developing larger study cohorts in non-European populations, would be more effective.

Differences in genetic effects across ancestries can be assessed by quantifying cross-ancestry genetic correlations (*ρ*_*g*_). A recent study^9^ estimated the genetic correlation of 31 complex traits between European and East Asian ancestries. The average genetic correlation was high (*ρ*_*g*_ = 0.85), suggesting that genetic effects on complex traits in European and East Asian populations are largely shared. However, the estimates varied substantially across different traits. For example, the genetic correlation estimates range from 0.34 for major depressive disorder to ∼1 for glomerular filtration rate (a measure of kidney function). In general, variable genetic correlation estimates across complex traits are expected because they are complicated by inconsistent disease phenotype definitions across studies, potential cell type composition differences in individuals of different ancestries^10–12^, environmental factors, gene-environment (GxE) interaction, and other non-genetic factors. Thus, it remains unclear how much genetic effects are shared across ancestries, and it is unknown whether differences in genetic effects compromise PRS portability.

Because gene expression can be measured quantitatively from the same cell type in a controlled environment, genetic correlation estimates among gene expression levels are expected to be less affected by non-genetic factors than correlation estimates for organism-level traits. Here, we studied the sharing of genetic effects on gene expression levels and the portability of gene expression level PRS between European and African individuals^13^. We elected to study gene expression in lymphoblastoid cell lines (LCLs), instead of organismal traits or gene expression in other cell types for several reasons. Firstly, gene expression levels have relatively sparse genetic architecture and large effects that imparts power to identify genetic effects^12,14^. Second, studying genetic effects in a single cell type allows us to avoid the complexity of cell-type specific *cis*-eQTL effects and the potential cell type composition differences in individuals of different ancestries^10–12^. Lastly and importantly, because these LCLs were cultured in the same lab environments, there is minimal environmental variation and contribution of GxE effects to gene expression differences between the two groups should be minimal. This experimental approach to minimize GxE improves our power to quantify the sharing of genetic effects and PRS portability.

Using gene expression data from LCLs derived from European and African individuals, we aim to answer a few fundamental questions. How do genetic effects differ between ancestries? How do LD differences, allele frequency differences and genetic effect differences across ancestries affect portability of gene expression PRS? And finally, how much do these differences explain differential gene expression between ancestries? The answers to these questions are essential for understanding and improving PRS portability.

By carefully analyzing gene expression levels in LCLs from European and African individuals, we provided concrete answers to all three questions for the first time. We found that *cis*-genetic effects are highly similar across ancestries, which agrees with a recent study in organism-level complex traits^15^ and is not expected to affect portability. Interestingly, we found that when causal variants differ in allele frequency between ancestries, PRS portability is substantially diminished independently of LD. Lastly, we found that allele frequency differences are also the major genetic factor underlying differential gene expression. Our findings significantly change the common understanding of PRS portability.

## Results

### *Cis*-genetic effects in European and African populations are highly similar

To compare genome-wide *cis*-genetic effects in different ancestral backgrounds, we obtained DNA- and RNA-sequencing data collected from LCLs as part of the GEUVADIS study^13^, which includes 358 individuals from four European (EUR) populations (CEU, FIN, GBR, and TSI; see Methods) and 87 Yoruba individuals from Ibadan, Nigeria (YRI). We used these samples to represent European and African populations, respectively. We performed *cis*-eQTL calling separately in each population, where *cis* regions are defined as regions within 100kb upstream and downstream from the transcription start sites. Our choice to focus on 200kb windows (instead of the 2Mb windows typically used for eQTL mapping^10,16^) allowed us to capture large effect *cis*-eQTLs while limiting the number of possible causal variants, which is useful in our downstream analysis. Using FastQTL^17^, we identified 2,885 genes with at least significant cis-eQTLS (ie. eGenes) in EUR and 469 eGenes in YRI that had at least one *cis*-eQTL at 10% FDR (Benjamini-Hochberg procedure). We estimated *cis* SNP heritability of gene expression in each population separately using GCTA^18^. Across all genes, the *cis* average SNP heritability is 0.119 in EUR (S.E.=0.003) and 0.096 in YRI (S.E.=0.002, Table S1).

To estimate the cross-population genetic correlation of *cis* effects on gene expression, we used the unconstrained bivariate GREML model from GCTA^18,19^. Across all genes, the average genetic correlation is 0.95 (S.E.= 0.09, **Figure 1A** and **1B**), which suggests that genetic effects on gene expression are largely shared between EUR and YRI populations. This result is surprising given that previous studies have reported much lower estimates of *cis-*genetic correlation between the two populations, for example, 0.313 in Brown et al^20^ and 0.47 in Mogil et al^21^).

**Figure 1.**
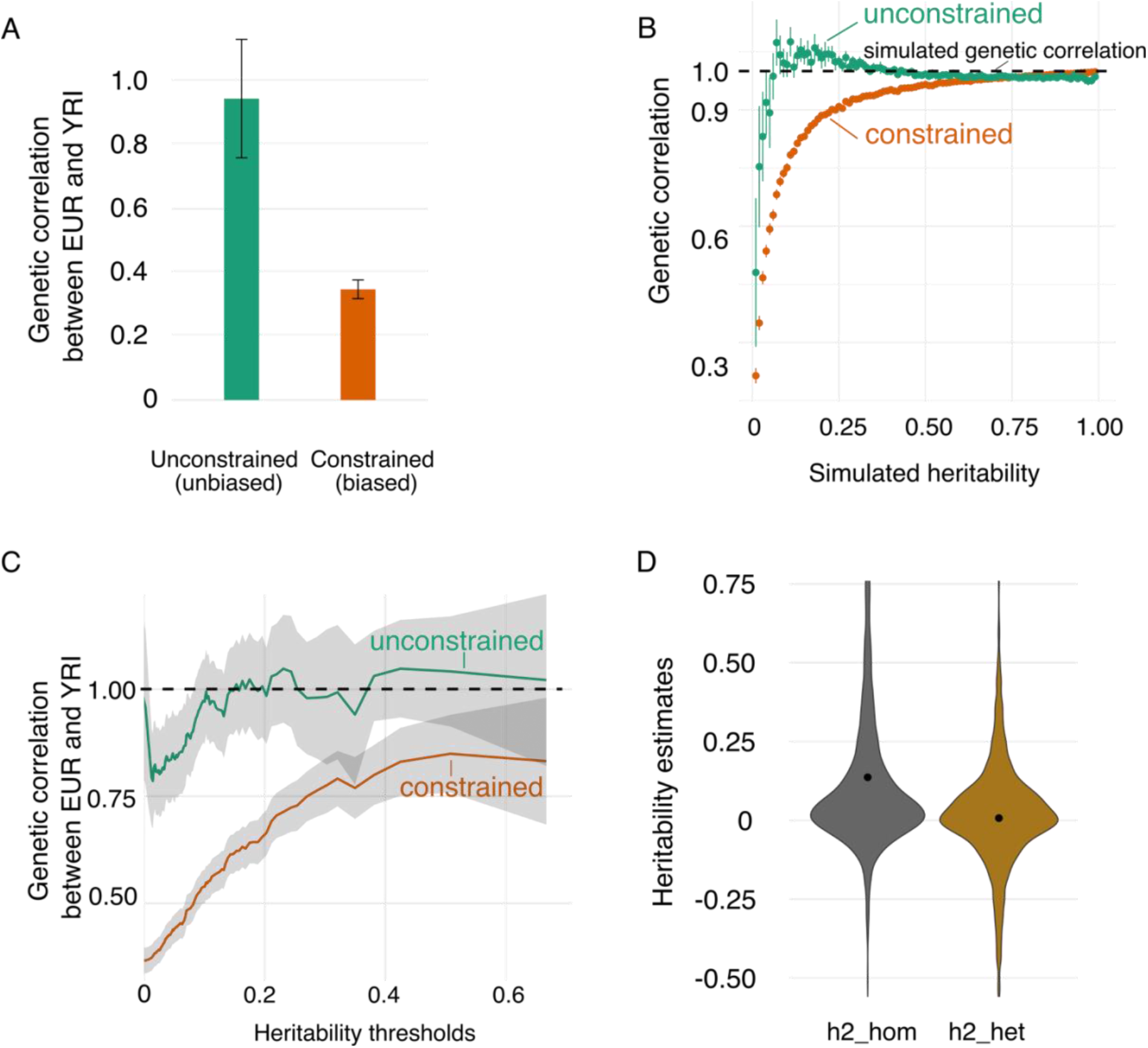
Highly shared genetic effects across ancestries. **A. Unbiased vs. biased genetic correlation estimates in GEUVADIS dataset**. Error bars are confidence intervals. **B. Unconstrained and constrained genetic correlation estimates in simulations**. The genetic correlation is set to 1 in all simulations. Unconstrained genetic correlation estimates (Green) are mostly unbiased and stable across different heritability settings. Constrained genetic correlation estimates (Orange) are substantially biased, especially at low heritability settings. Error bars are confidence intervals. **C. Genetic correlation estimates at different heritability thresholds in GEUVADIS dataset**. Y-axis is the average genetic correlation estimates for all genes with estimated heritability larger than a heritability threshold (defined in X-axis). Constrained genetic correlation estimates (Orange) are much lower than unconstrained genetic correlation estimates, which agrees with the simulation results. Constrained estimates show a pattern of increasing genetic correlation at higher heritability thresholds, which is a thresholding bias also seen in ref^20^ Figure 3 and ref^21^ Figure 2. **D. Heritability explained by shared and population-specific effects across ancestry**. Heritability explained by population-specific effects across ancestries is h2_het (Orange). Heritability explained by shared effects across ancestries is h2_hom (Orange). Confidence intervals are plotted as error bars but are too small to be visible.

To better understand the cause of this discrepancy, we evaluated the methods employed by previous studies. Heritability estimates are typically constrained to a range from 0 to 1, and genetic correlation estimates are usually constrained to a range from -1 to 1 (including ref^20^ and ref^21^). However, we previously showed that constraining heritability between 0 and 1 can produce substantially biased heritability estimates^22^. Here, we show that the same principle applies to constraining genetic correlation estimates. To demonstrate this bias, we simulated genetic effects on gene expression using real EUR and YRI genotypes within *cis* regions in GEUVADIS. Specifically, we simulated gene expression levels assuming various levels of SNP heritability (h^2^_g_) and identical effects in each ancestry (*ρ*_*g*_ = 1) (see Methods for details). We used GCTA to estimate both the standard constrained genetic correlation estimates (i.e. constrained from -1 to 1, as was done in previous studies) and the unconstrained estimates. We performed 10,000 simulations for different h^2^ values ranging from 0 to 1, totaling nearly 1,000,000 simulations. We found that constraining the genetic correlation estimates caused substantial downward biases, especially at lower simulated heritability levels (**Figure 1B**, Figure S1). In contrast, unconstrained estimates are largely unbiased and much more stable across almost all heritability values. We also confirmed empirically that constraining the genetic correlation estimates in our dataset resulted in a substantially lower average genetic correlation of 0.35 (S.E.=0.01) (**Figure 1A**) and lower genetic correlation estimates across all heritability bins (**Figure 1C**).

To further examine the sharing of *cis* genetic effects across ancestries, we used a mixed model implemented in GxEMM to quantify the proportion of gene expression heritability explained by shared and population-specific genetic effects^23^ (see Methods). Across all genes, the proportion of heritability explained by shared (h2_hom) effects is 0.136 (S.E.=0.006). In contrast, the proportion of heritability explained by ancestry-specific (h2_het) genetic effects is 0.007 (S.E.=0.004, P value=0.1, **Figure 1D**, Table S1), which is not significantly greater than 0 and indicates that population-specific effects explain a negligible proportion of gene expression heritability. These results are expected based on our cross-ancestry genetic correlation estimates being indistinguishable from 1 and further emphasize that genetic effects on gene expression are highly shared between EUR and YRI.

### PRS portability is significantly impacted by allele frequency differences at causal loci

We next studied the portability of gene expression prediction (gene expression PRS) across ancestries in the GEUVADIS data. We split the 358 EUR samples into 271 training samples and 87 prediction samples. We trained SNP weights of *cis* genetic variants using elastic-net in 271 EUR samples; we then used the trained weights to predict gene expression in 87 YRI and 87 held-out EUR samples. We defined portability from EUR to YRI as the ratio of prediction R^2^ in YRI to the prediction R^2^ in EUR. Across genes, we estimated the portability as 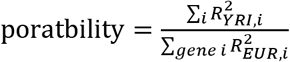. This ratio of the averages is more stable than the average of the ratios. We obtained standard errors (S.E.) of portability by jackknifing across genes. We estimated that the average portability is 0.63 (S.E.=0.01) across the 7360 genes, which has at least one SNP with non-zero coefficient fitted by elastic-net in the EUR samples. We note that these genes are likely a subset of genes with high heritability in EUR samples. Therefore, the portability estimates of these genes do not represent genome-wide estimate of portability, because portability depends on heritability in EUR (Figure S2). Yet this approach accurately reflects common practice of PRS construction and validation, in which PRS typically has reasonable accuracy in EUR. The prediction accuracy R2 and portability of genes can be found in Table S2.

We found that the portability of gene expression PRS was highly variable across genes. This variability allowed us to examine the effects of different genetic factors on PRS portability. Since we found that genetic effects are highly shared between EUR and YRI, population specific effects are not expected to contribute much to portability. We therefore examined how differences in LD patterns within *cis* regions across ancestries and differences in causal *cis* variant allele frequencies could impact portability (**Figure 2**). We stratified genes based on LD pattern differences in *cis* regions across the two populations. We quantified the extent of cross-ancestry LD differences in *cis* regions by using varLD^24,25^. The varLD method first performs eigen decomposition on the local LD matrices for each population and then uses the differences in the eigenvalues as varLD scores ^24^. Higher varLD scores indicate larger LD differences (see Methods, Table S3). We observed high portability (72%, S.E.=0.11, **Figure 2A**) when LD patterns are highly similar in EUR and YRI (i.e., the lowest 5 percentiles of varLD scores). PRS portability decreases with increasing dissimilarity in LD, and the portability is only 54% (S.E.=0.11) at the highest 5% of varLD scores. These observations show that LD differences influence portability.

**Figure 2.**
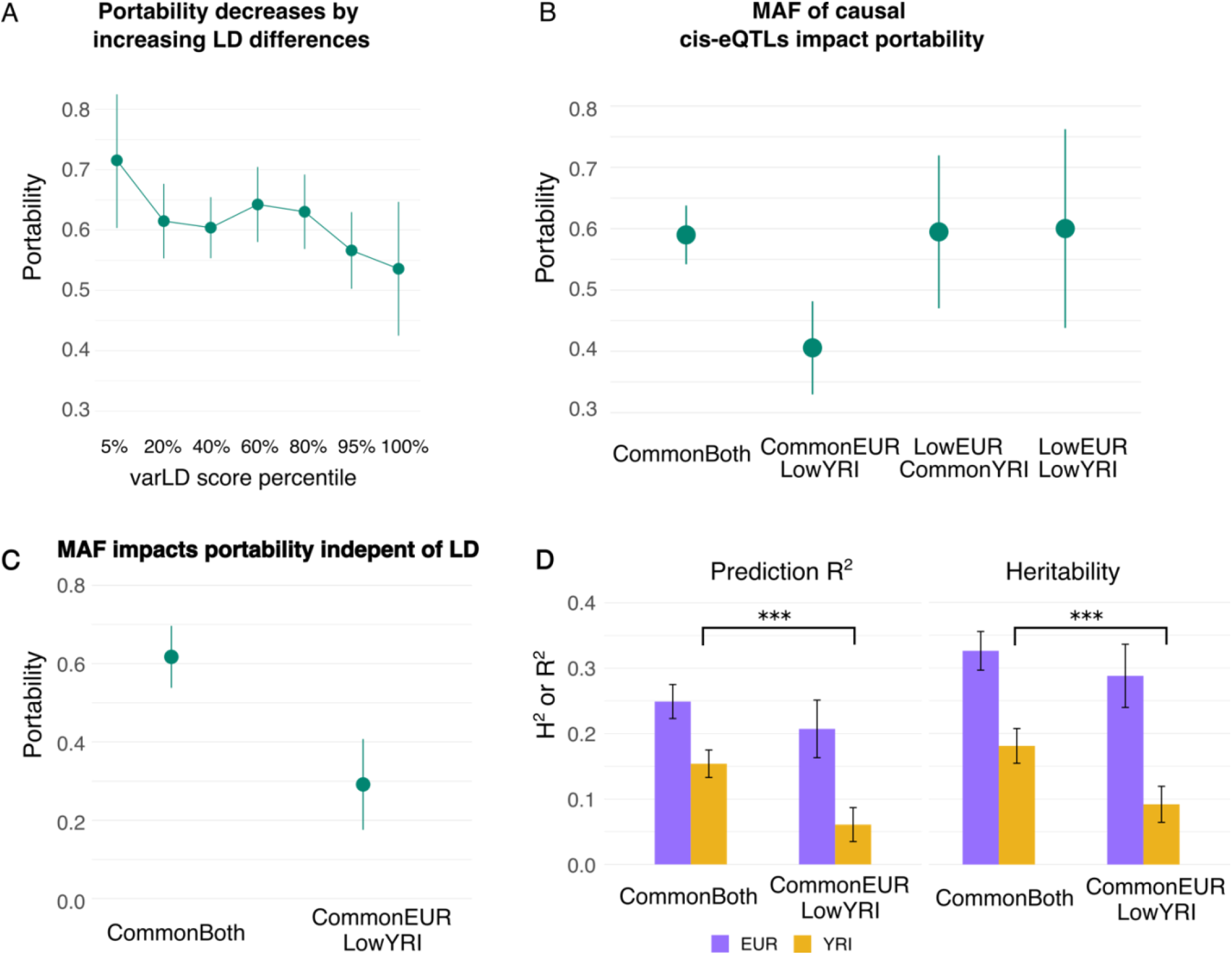
Effects of different genetic factors on portability. **A. Portability of genetic scores of gene expression decreases by increasing LD differences**. The sharing of LD patterns of *cis* regions were quantified by varLD scores, where smaller varLD scores indicate smaller differences in LD and larger varLD scores indicate larger differences in LD. Error bars are confidence intervals. **B. Portability from EUR to YRI drops significantly in genes whose causal cis-eQTLs are common in EUR but low in YRI**. Genes are stratified by the allele frequency of causal cis-eQTLs fine-mapped in GTEx LCL samples^25^. Low frequency variants are variants with MAF < 5%, whereas common variants are variants with MAF > 5%. Error bars are confidence intervals. **C. Portability of a subset of 192 genes whose causal cis-eQTLs are used to predict gene expression levels**. Error bars are confidence intervals. **D. Heritability and portability of 192 genes whose causal cis-eQTLs are included in predicting PRS**. Y axis on the left are SNP heritability and prediction accuracy measured in R^2^. Y axis on the right is portability. Error bars are confidence intervals. The stars denote the significance of differences in prediction accuracy R^2^ and heritability. P-value of difference (t-test) for prediction accuracy R^2^ in YRI is 2.25×10^−7^; P-value of difference (t-test) for heritability in YRI is 8.30×10^−6^.

To demonstrate the impact of causal allele frequencies on portability, we stratified genes by the allele frequencies of the fine-mapped *cis*-eQTLs. To avoid winner’s curse, we used fine-mapped *cis*-eQTLs identified in GTEx LCL samples ^25^, which do not overlap with our GEUVADIS samples. We used fine-mapped SNPs from Wang et al^25^, which performed fine-mapping of *cis*-eQTLs by applying SuSiE^26^ with functionally informed priors, which improve accuracy over flat priors. For each gene, we define its causal *cis*-eQTL as the fine-mapped *cis* variant with the maximum posterior inclusion probability (PIP) in the GTEx LCL samples. We focused on a subset of 1025 genes with fine-mapped *cis*-eQTLs and portability estimates in our study. We stratified these genes into categories based on the GEUVADIS allele frequencies of the causal *cis*-eQTLs: causal *cis*-eQTLs common (MAF>5%) in both populations (CommonBoth), causal *cis*-eQTLs common in EUR but low frequency in YRI (commonEUR-lowYRI), and causal *cis*-eQTLs common in YRI but low frequency in EUR (lowEUR-commonYRI). We observed that the PRS portability of genes in the CommonBoth category is 0.59 (S.E.=0.05, **Figure 2B**). Interestingly and importantly, we observed a drop in PRS portability of **more than 32%** (P value of difference is 2.9×10^−3^) for genes in the commonEUR-rareYRI category, whose average portability is only 0.40 (S.E.=0.08; **Figure 2B**). These results support that allele frequency differences of causal *cis*-eQTLs have a significant impact on PRS portability.

It is difficult to tease apart the impact on portability from LD versus causal allele frequencies in studies of organismal complex traits, because of the highly polygenic genetic architecture and small genetic effects at the majority of disease loci. In our study, we leveraged high quality fine-mapped *cis*-eQTLs to separate the impact of LD and causal MAF on portability. To demonstrate that causal allele frequency differences can significantly impact portability independent of LD, we focus on a subset of 192 genes in the CommonBoth and CommonEUR-LowYRI categories, whose causal variants were successfully included in PRS. In this set of genes, LD is expected to contribute little to prediction portability, and yet we observed much lower portability in the CommonEUR-LowYRI category (**Figure 2C**, Methods) than in the CommonBoth category (P value of difference=8.5×10^−4^). The substantial decrease in portability in the CommonEUR-LowYRI category can be traced to the low heritability and prediction accuracy explained by low frequency/rare causal variants in YRI (**Figure 2D**). More specifically, we observed significantly lower prediction accuracy R^2^ and heritability of genes in YRI of the CommonEUR-LowYRI category than the CommonBoth category (P-value of difference for prediction accuracy R^2^=2.25×10^−7^; P-value of difference for heritability=8.30×10^−6^; t-test), while the heritability in EUR is comparable across the two categories (P-value of difference for prediction accuracy R^2^=0.10; P-value of difference for heritability=0.18; t-test, **Figure 2D**).

Finally, we examined how often genes have different causal allele frequencies and are therefore expected to have low portability. In the set of 1,851 genes whose *cis*-eQTLs were successfully fine-mapped in GTEx LCLs^25^, the allele frequencies of causal *cis*-eQTLs are largely comparable across EUR and YRI (Pearson correlation = 0.66, p value<10^−16^, Figure S3A). However, we observed a slightly higher proportion of low frequency variants in YRI (Figure S3B and S3C), likely because the GTEx dataset is predominantly of European ancestry. 288 (15.6%) genes have causal *cis*-eQTLs that are common in EUR but rare in YRI, which are likely to suffer from low PRS portability. We observed a similar proportion in whole blood eQTL data from GTEx: 1,097 out of 6,392 (17.2%) genes that were successfully fine-mapped^25^ have causal *cis*-eQTLs that are common in EUR but rare in YRI.

### Allele frequency difference of causal variants and non-genetic effects drive differential expression across ancestries

Many complex traits and diseases vary across different ancestral groups. Organismal-level phenotypic differences, including differences in disease prevalence, may be mediated by differential gene expression. Many genes are differentially expressed across ancestries, but the mechanisms driving differential expression are largely unknown.

To better understand what drives differential expression across populations, we performed differential gene expression analysis in the GEUVADIS dataset. We identified 6,128 differentially expressed genes (FDR < 5% and absolute expression fold change > 1.1, Table S4) using DESeq2^27^. To identify causal *cis*-eQTLs, we used SuSiE^26^ to fine-map causal variants in EUR and YRI populations separately. We identified 95% credible sets of causal variants for 2,096 genes in EUR and 347 genes in YRI (Methods). We used the overlap of variants in credible sets as a proxy for shared common causal effects. However, we note that due to incomplete power, overlap does not necessarily imply sharing of the causal variants themselves. Similarly, a lack of overlap does not imply the existence of population-specific causal variants. We fine-mapped a total of 190 genes to credible sets with at least one overlapping variant in YRI and EUR. We defined the causal *cis*-eQTL of each gene to be the SNP with the maximum product of PIPs in the two populations.

We first examined the impact of causal *cis*-eQTL allele frequency differences on differential gene expression. Among the fine-mapped SNPs for each eGene, we defined the expression-increasing allele as the allele that has positive effects in both EUR and YRI populations. We found that allele frequency differences strongly correlated with gene expression differences (**Figure 3A**, Table S5). Specifically, for genes exhibiting higher expression levels in EUR (purple dots, “higher EUR” in **Figure 3A**), the allele frequencies of expression-increasing alleles are significantly higher in EUR (Pearson correlation=0.4; p value=0.0058). This pattern also holds for genes exhibiting higher expression levels in YRI (Pearson correlation=0.4, p-value =0.01; orange dots in **Figure 3A**) and the genes without significant differential expression (Pearson correlation=0.32, p-value=0.0051, gray dots in **Figure 3A**). For example, the *cis*-eQTLs of *PPIL3* were fine-mapped to rs7559150 in both populations, but *PPIL3* exhibits higher expression in EUR (**Figure 3B**). As expected, the expression-increasing T allele has a much higher allele frequency in EUR (0.736) than in YRI (0.259) (**Figure 3B**).

**Figure 3.**
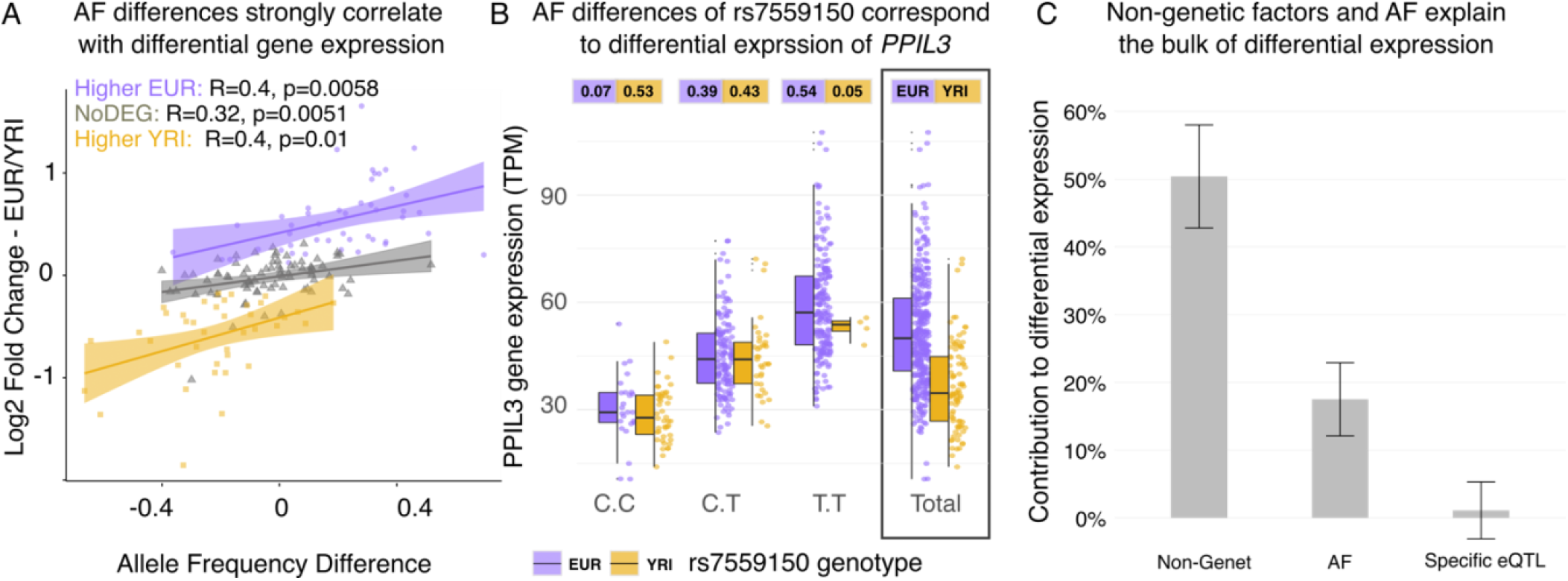
Cross-ancestry differential gene expression can be explained by allele frequency (AF) differences and non-genetic factors. A. Allele frequency differences of the expression-increasing allele strongly correlate with gene expression fold change in the two populations. Purple dots denote *cis* variants of genes that are significantly more expressed on EUR than in YRI. Orange dots denote cis-variants of genes with significantly higher gene expression in YRI than in EUR. The lines are the lines of best fit. Gray dots and the gray line denote variants of genes that are not significantly differentially expressed between the two populations. Allele frequencies are the frequencies of the expression increasing allele (Table S5). **B. Allele frequency differences of rs7559150 correspond to differential expression of *PPIL3***. Purple denotes the expression levels of the European individuals carrying CC/CT/TT genotypes and the overall expression levels. Orange denotes the expression levels of the Yoruba individuals carrying CC/CT/TT genotypes and the overall expression levels. The numbers in purple and orange boxes are the genotype frequencies in EUR and YRI samples. **C. The proportion of differential gene expression explained by non-genetic factors, allele frequency difference, and population-specific genetic effects**. Non-Genetic factor and AF explain bulk of differential expression, while population-specific effects explain little differential gene expression. Error bars are confidence intervals.

We next sought to quantify how allele frequencies and eQTL effects contribute to differential gene expression between populations. We developed a model to partition differential gene expression into three terms: (1) population-specific eQTLs; (2) population AF differences (3) nongenetic population differences (Methods, Supplement). Component (1) captures differences in population-specific (non-portable) causal effects. Component (2) captures population differences in allele frequency at eQTLs. Component (3) captures both nongenetic differences as well as unmodelled genetic effects in *cis* or trans. No matter how large the sample size from population A, the information in components (1) and (2) will never benefit prediction in population B; in contrast, the nongenetic factors in component (3) are irrelevant for PRS portability, specifically, though they could certainly be relevant for other questions.

We applied this model to 190 genes where SuSiE identifies an overlapping high-confidence fine-mapped eQTL in both populations (**Figure 3C**). On average, population-specific eQTL effects explain 1.1% (S.E.=2.1%) of differential gene expression, which is not significantly different from 0. AF differences explain 17.5% (S.E.=2.7%) and non-genetic differences explain 50.4% (S.E.=3.8%). Thus, we conclude that in this set of well-mapped eQTLs, AF differences are the dominant genetic component, and non-genetic differences are the major player overall.

## Discussion

Our study aimed to answer three important questions: How similar are genetic effects on gene expression across ancestries? How do differences in causal allele frequencies, causal effect sizes, and LD patterns affect PRS portability? What factors drive differential gene expression across populations? By carefully analyzing gene expression in LCLs from EUR and YRI individuals, we provided concrete answers for all three questions for the first time.

We found that *cis* genetic effects on gene expression in LCLs are highly shared across between EUR and YRI populations. This contrasts with previous low estimates of genetic correlation, which were proved to be statistical artifacts from constraining correlation estimates. This bias is large when sample size and/or heritability are low, but is not likely to substantially affect analyses in large datasets^28^. A caveat is that our approach does not model population-specific LD patterns; nonetheless, this is unlikely to upwardly bias our estimates, hence our results are conservative. While we observed high average genetic correlation in gene expression, we stress that this will not necessarily be the case for all complex traits. The etiology of organism-level complex traits may involve multiple cell types, tissues and their interactions, which largely remain unclear. Organism-level complex traits may also have higher contributions of environmental factors, and, likely, gene-environment interactions, both of which may vary across individual populations. Finally, we did not model epistasis. A recent study has shown that epistasis can drive heterogeneity of causal effects for gene expression and other complex traits^29^, although it is unclear if epistasis is important for complex traits.

We found that allele frequencies of causal *cis*-eQTLs play an important role in reducing portability of gene expression prediction. When causal variants are common in EUR but rare or low frequency in YRI, we observed a ∼32% drop in portability. Causal allele frequency can impact PRS portability through the influence of LD, and it is difficult to tease apart LD and causal allele frequency differences. In our analyses, we managed to tease apart the impact of LD and causal allele frequency differences on portability for the first time by using carefully fine-mapped SNPs to identify causal variants with large effects. We focused on a subset of genes whose causal variants are included in the PRS variants to predict gene expression levels. We found that allele frequency difference, separately from LD, can significantly impact portability. A direct implication of this observation is that even when causal variants were successfully fine-mapped, portability would remain poor if the causal variants have low or rare allele frequency in the PRS population samples. Therefore, improved statistical fine-mapping alone is not sufficient to improve PRS portability caused by causal allele frequency differences; instead, equitable PRS prediction requires novel methodological approaches and increased diversity in GWAS data.

We found that a sizable fraction (15.6%-17.2%) of genes have causal *cis*-eQTLs that are common in EUR but have low frequency in YRI, and these genes are expected to suffer low portability. Large allele frequency differences between populations of different ancestries are common because of genetic drift, demographic histories and, in some cases, natural selection. We expect that the impact of frequency differences on portability to vary across different complex traits, because they are under different levels of selection. For example, recent studies have shown that complex traits under strong negative selection suffer worse portability^30^. Organismal complex traits undergo different levels of selection pressure than gene expression levels, and the impact of causal allele frequency difference on portability of organismal level complex traits should be further evaluated.

There are several limitations of the study. First, we assumed that there was one causal variant in 200kb cis-regions of each gene in statistical fine-mapping, which may not hold true for every gene. However, recent study^12^ has shown that more than 80% eGenes in GTEx LCL have only one *cis*-eQTL in the 2Mb *cis-*region. Our definition of cis-region is much shorter than 1Mb and the one *cis*-eQTL per gene assumption might work well for most genes. Second, the tools we used to estimate heritability and genetic correlations, such as GCTA, work the best with polygenic or even infinitesimal genetic architecture, yet *cis* gene regulation is known to have a sparse genetic architecture. Our previous has shown that GCTA generates unbiased results in very sparse settings^31^. We also performed simulation with GCTA where only 1% of SNPs in the 200kb *cis* regions are causal, and unbiased estimates were generated under the unconstrained model for most heritability settings (Figure 1B). Third, we assumed that the fine-mapped causal variants by SuSiE^26^ are the causal variants in our analyses of portability and differential gene expression. SuSiE is among the best methods for fine-mapping causal variants. While the fine-mapping may not be perfect, we demonstrated that the prediction accuracy and the heritability of genes is higher when causal allele frequency is common in YRI and lower when the allele frequency is of low frequency, which are expected when we have the true causal variants. We also showed that the causal allele frequency correlates well with the differential gene expression levels, which is also expected when we have the true causal variants.

Finally, while there are other gene expression datasets involving samples of multiple ancestries, GEUVADIS provided the best available dataset for our analyses, despite its small sample sizes. Other gene expression datasets^21,32^ are either comprised of microarray data that are significantly noisier than RNA-sequencing datasets (average *cis* SNP heritability is only 0.02 in array data at much larger sample sizes in comparison to 0.1 in RNA-seq data^33,34^), or the datasets contain admixed samples that require more complex statistical methods, which should be developed in future work. Future datasets with larger sample sizes, more complex traits and a more equitable collection of continental ancestries will further improve our understanding of the extent to which genetic effects are shared across ancestries and, in turn, improve the clinical utility of PRS for all.

## Materials and Methods

### Gene expression dataset

We used the transcriptome data of the 445 unrelated individuals from the GEUVADIS project^13^, where LCL RNA-sequencing data are collected from African (Yoruba in Ibadan, Nigeria, or YRI) and European groups (https://www.ebi.ac.uk/arrayexpress/files/E-GEUV-1/). We validated the sequencing quality of the fastq files using FastQC (http://www.bioinformatics.babraham.ac.uk/projects/fastqc/). The RNA-sequencing reads were mapped to the human reference genome (hg19) and quantified using Kallisto ^35^. Only 12980 genes with counts per million reads (CPM) > 0.5 in more than half of the total samples were kept for further analysis. Expression levels of these genes were quantified as Transcripts Per Million (TPM). We performed quantile normalization across all samples and then normalized the expression levels to a standard normal distribution across genes. We used the SVA^36,37^ package to identify surrogate variables. We excluded 235 genes in the HLA region (hg19 chr6: 29691116–33054976) due to their extreme variation and complexity.

### Genotype dataset

The genotypes of GEUVADIS samples are in the 1000 Genomes phase 3 dataset. Of the 465 unrelated individuals in the GEUVADIS dataset, we used 445 matched individuals in the 1000 Genomes genotype data (358 European and 87 Yoruba individuals)^13^. Using VCFtools^38^, we selected bi-allelic autosomal variants with >1% minor allele frequency in the total population for the subsequent eQTL analysis. Allele frequency and genetic principal components (PCs) were calculated by plink 1.90 ^39^ and VCFtools^38^. We excluded 44 multi-allelic SNPs that are reported as bi-allelic SNPs.

### *Cis*-eQTL mapping analysis

We used fastQTL ^17^ for *cis*-eQTL mapping. We defined cis-region as 100kb upstream and downstream regions from the transcription start site (TSS) for each gene. Sex, the top three gene PCs, and seven surrogate variables were used as covariates. Permutation analysis was conducted with a total of 1,000 permutations^17^. In total, we identified 2885 *cis*-eGenes with at least one *cis*-eQTL in European and 469 *cis*-eGenes with at least one *cis-*eQTL in Yoruba, at 10% FDR (Benjamini-Hochberg procedure) in each population.

### Genetic correlation analysis and simulations

We used the bivariate genome-based restricted maximum likelihood (GREML) analysis in GCTA ^18,19^ to estimate the genetic correlation of gene expression between EUR and YRI samples in GEUVADIS. By default, GCTA constrains genetic correlation estimates to lie between -1 and 1. We also fit unconstrained genetic correlation estimates using the option “--reml-no-constrain --reml-bivar-no-constrain” option when running GCTA. Without this command, GCTA wil yield genetic correlation estimates constrained to a range between -1 and 1.

We performed extensive simulations to evaluate biases of genetic correlation estimates. We used real genotypes (denoted as X) of *cis*-regions from a randomly selected gene in GEUVADIS EUR and YRI samples. We simulated identical genetic effects in EUR and YRI, *ie. ρ*_*g*_ = 1. We simulated genetic effects, β, by assuming p=1% of SNPs in the *cis* region are causal. Specifically, β follows a point normal distribution, such that 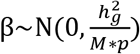 for p=0.01, and β = 0 otherwise. 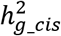 is the *cis* heritability, which is set to a sequence of 100 values between 0 and 1 (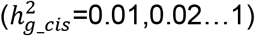). M is the number of SNPs in the *cis* region. Expression y in each population was simulated as y = Xβ + ϵ, where 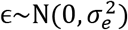. We performed 10,000 simulations at each heritability value, totaling 1,000,000 simulations. We also performed 1,000,000 simulations assuming p=10%, and the results can be found in Figure S1.

For each simulation, we performed bivariate GREML analysis to generate the unconstrained and constrained genetic correlation estimates. We computed the average genetic correlation estimates at each *cis* heritability level.

### Estimating heritability of gene expression explained by population specific effects

To quantify gene expression heritability explained by shared and population specific effects, we used GxEMM^23^. GxEMM uses a mixed model to estimate the heritability specific to different environments (h2_het) and heritability shared by the environments (h2_hom). We used binary population labels as the “Environments” (E) in GxEMM. We fit GxEMM using the HE regression option because it is computationally stable and roughly unbiased estimates, even for small sample sizes.

### Genetic prediction of gene expression and portability between EUR and YRI populations

To compute genetic scores of gene expression (gene expression PRS) and the portability of genetic scores, we first split the EUR samples from the GEUVADIS dataset into 271 training samples and 89 testing samples. The 89 YRI samples were also used as testing samples. We used the glmnet R package^40^ to fit the elastic net model to predict gene expression of the 271 training samples from *cis* region SNP genotypes. We used the mixing parameter of 0.5, and we also included three genetic PCs and seven surrogate variables as the covariates in the model. We then used the SNP weights to derive the PRS of gene expression in the 89 EUR and 89 YRI samples, by computing the weighted sum of genotypes. The *R*^2^ of PRS in each population were computed with *R*^2^(actual gene expression, predicted PRS). We defined the portability as 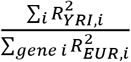.

### VarLD Analysis

We quantified the LD differences of *cis* regions between EUR and YRI using varLD^24,41^. VarLD first quantified LD using signed r^2^ between every pair of SNPs in a predefined window, which resulted in a symmetric LD matrix in each population. Eigen decomposition was then performed on the LD matrices. Raw varLD scores were computed as the sum of the absolute differences between the ranked eigenvalues of the two populations. In our analysis, each window was defined as 200 SNPs common to the two populations, as suggested in ref^24^. We ran a sliding window of 200 SNPs in the *cis* regions of all genes. We normalized the raw varLD scores across all the windows. Finally, for each gene, we used the maximum varLD scores within the sliding windows to represent the varLD score of the *cis* region. The maximum varLD score for each gene can be found in Table S3.

### Differentially expressed gene analysis

Differential expression analysis was performed by DESeq2^27^, which calculates the fold change of transcription of each gene using the Wald test and a correction for multiple hypotheses. Sex, the top three genetic PCs, and seven surrogate variables were used as covariates. We defined genes with an adjusted p-value of < 0.1 and absolute expression fold change > 1.1 as differentially expressed between the two populations. We identified 6,128 differentially expressed genes using DESeq2.

### Fine-mapping *cis*-eQTLs

To fine-map causal *cis*-eQTLs, we used SuSiE^26^. We define *cis* regions as 100kb upstream and downstream regions from the transcription start site of each gene. We then ran susieR on the *cis*-eGenes for each population separately, allowing up to 10 credible sets (L = 10, coverage = 0.95, scaled_prior_variance = 0.95). 95% credible sets of *cis*-eGenes were reported in both populations. For each gene, if the credible sets have at least one overlapping SNP, we define the overlapping SNPs as the “shared” causal SNPs. We also tried using EMS as priors to fine-map causal *cis*-eQTLs in GEUVADIS data. However, since EMS priors resulted in smaller credible sets, there were only 23 genes with “shared” causal SNPs, which is too few genes for downstream analysis. Therefore, we only used the susieR results of flat priors in our main analysis.

### Model to partition differential gene expression

To partition differential gene expression into different components, we assume a simple additive model for gene expression, assuming a single shared causal *cis*-variant per gene:

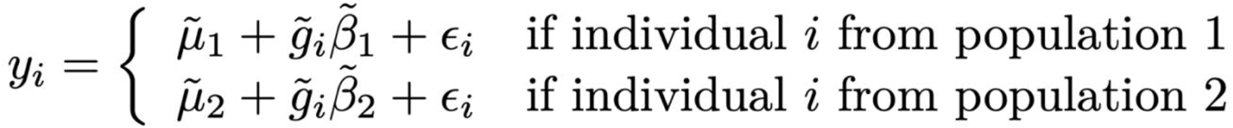

where *y* is the expression of a gene. 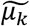 is the population-specific mean expression. 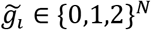 is the genotype of the causal SNP. 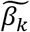 is the eQTL effect size in population k, and *ε*_i_ is random noise.

We define genotype mean in population *k, α*_k_ as: 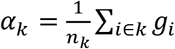. We then decompose the causal effect size by : 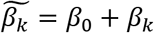, where : *β*_0_ is the population-shared eQTL effect and *β*_k_ is the population-specific eQTL effect. Likewise, we decompose the genotype into population-specific variation by 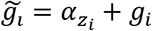, where *z*_*i*_ is an indicator whether sample *i* is in population 1 or 2.

Using these definitions, we can rewrite the above model in terms of population-shared and -specific parameters. This allows us to decompose differential gene expression as:

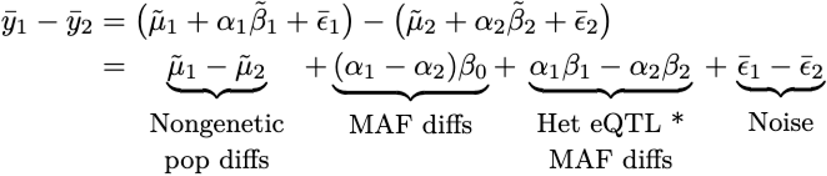

The first term captures mean differential expression between populations due to nongenetic effects and/or unmodelled eQTLs. The second term captures the effect of population-level allele frequency differences (*α*_*k*_), which is scaled by the shared eQTL effect size in each population. The third term capture population-specific eQTLs, which are weighted by their respective MAFs. The fourth term is the contribution of noise. The key feature of this decomposition is that it is invariant to recoding the reference allele for SNP *g*--because the sign of *α*_*k*_ flips and so do the signs of *β*_0_ and *β*_*k*_—and to changes in the overall MAF or nongenetic mean—because only the differences between populations enter the equation. We note that generalizing this approach to multiple populations (and/or admixed populations) is mathematically straightforward, but the notation is considerably more complex.

### Partitioning GEUVADIS eQTLs into population-shared eQTLs

For each gene that has overlapping credible sets between EUR and YRI, we obtain the shared fine-mapped set of *cis*-variants. Since we assumed there was only one causal variant in the *cis* region, we used only the top fine-mapped variant per gene, defined as the variant with the greatest product of population-specific PIPs. We partitioned the variance of gene expression into four components: non-eQTL population differences; MAF differences; specific eQTL effects, and noise. For each gene, we calculate the proportion of variance explained across these four components. We also adjust for the same technical covariates as used in fine-mapping, but we exclude this component from our decomposition.

## Data Availability

https://github.com/XuanyaoLiuLab/AF_PRS_portability

## URLs

All codes used in this analysis can be found in https://github.com/XuanyaoLiuLab/AF_PRS_portability

## Acknowledgements

We thank Yang Li for useful discussion and feedback. We thank Taro Kunimitsu for his support in coding. We thank Natalia Gonzales, Christian Jones, Sarah Sumner and Torgeir R Hvidsten for editing the manuscript.

All the analyses are done at the University of Chicago’s Research Computing Center.

## Notes

### Competing Interest Statement

The authors have declared no competing interest.

### Funding Statement

NIGMS

### Author Declarations

The GEUVADIS dataset is available at https://www.ebi.ac.uk/biostudies/arrayexpress/studies/E-GEUV-4

