## Supplementary Figures S1-S3 for "Allele frequency differences of causal variants have a major impact on low cross-ancestry portability of PRS"

*The authors contributed equally.

+Corresponding author.

### Supplementary Notes

Supplementary Figures

**Figure S1. Unconstrained and constrained genetic correlation estimates in simulations.**

**Figure S2. Portability depends on heritability in EUR. Genes are stratified by the heritability in EUR. Y axis is the prediction accuracy R2. Portability is the ratio of R2 between AFR and EUR.**

**Figure S3. Allele frequencies of causal cis-eQTLs inferred in GTEx LCL in EUR and YRI.**

Supplementary Tables

**Table S1 Genetic correlation, heritability, h2_hom and h2_het of genes**

**Table S2 R2 and portability of genetic scores of all investigated genes**

**Table S3 Max varLD scores of all investigated genes**

**Table S4 Differential expressed genes**

**Table S5 Frequency of the expression-increasing allele of the fine-mapped variants in EUR and YRI**

### Supplementary Figures

**Figure S1. Unconstrained and constrained genetic correlation estimates in simulations.** We performed simulations to evaluate biases of genetic correlation estimates. We simulated identical genetic effects in EUR and YRI, *ie.* $\rho g=1$. We assumed 1% or 10% of SNPs in cis region are causal, ie. p=1%. We simulated a sequence cis heritability values ranging from 0 to 1 (=0.01,0.02…1). We performed 10,000 simulations at each heritability value, totaling 1,000,000 simulations. The figure below shows the simulation results at p=10%. The simulation results for p=1% can be found in Figure 1B.

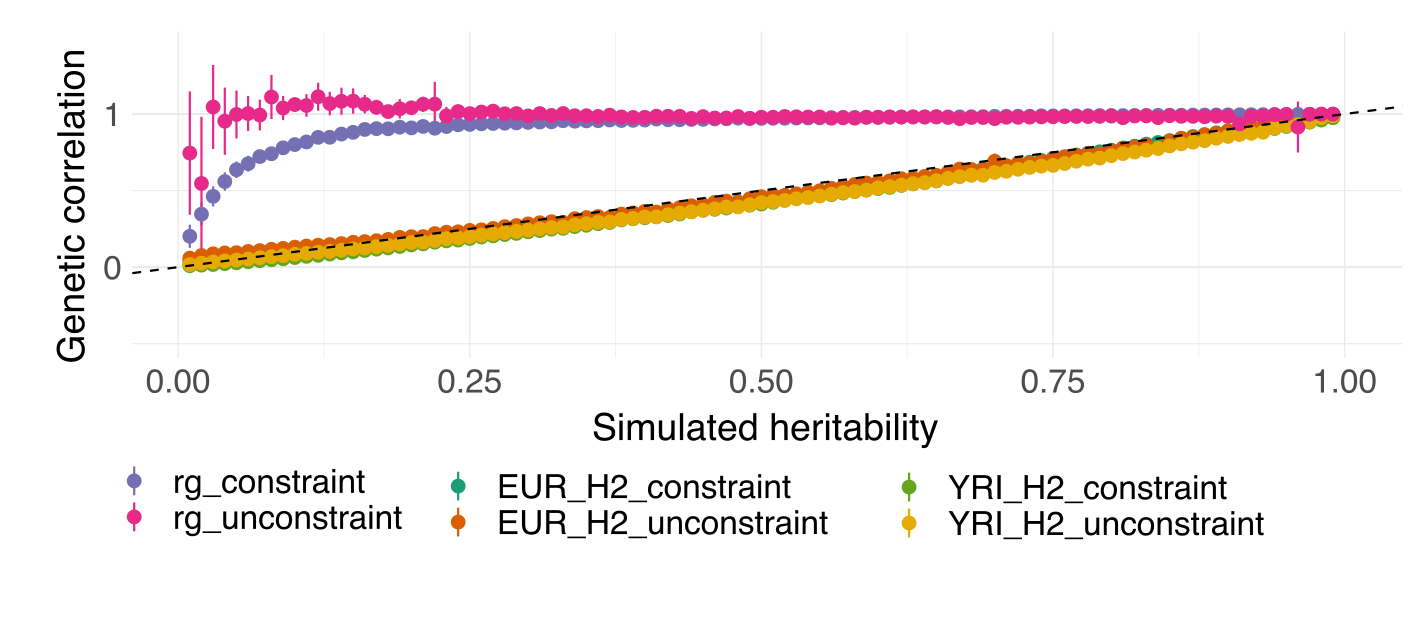

**Figure S2. Portability depends on heritability in EUR. Genes are stratified by the heritability in EUR. Y axis is the prediction accuracy R2. Portability is the ratio of R2 between AFR and EUR.**

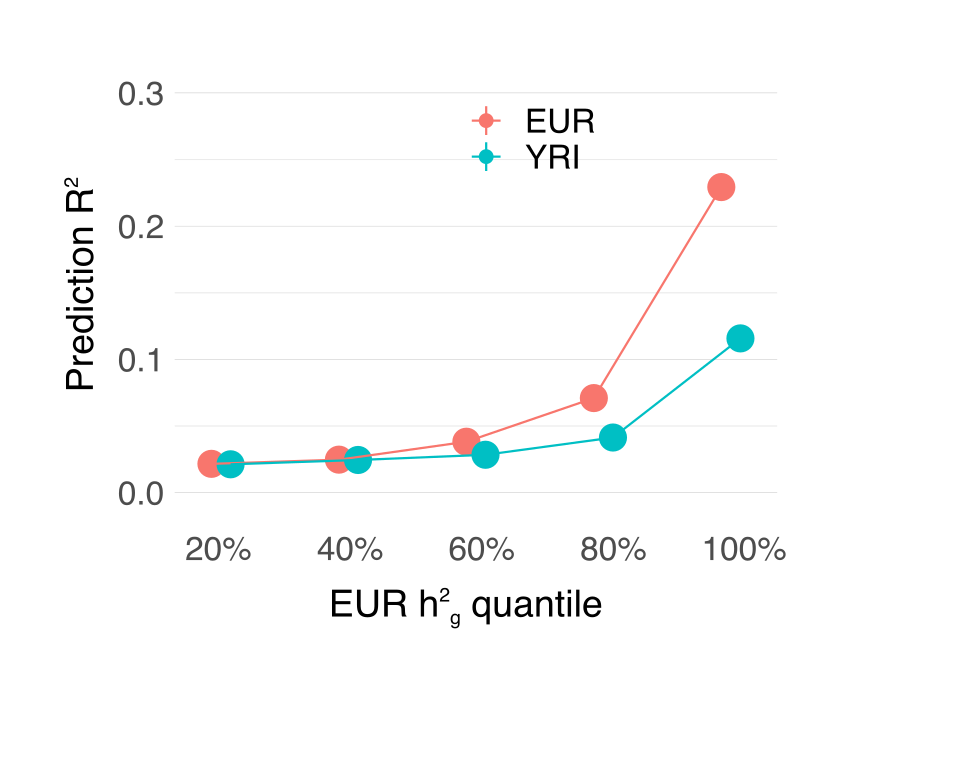

**Figure S3. Allele frequencies of causal cis-eQTLs inferred in GTEx LCL in EUR and YRI. (A) Scatter plot of causal allele frequencies in EUR vs YRI (B) Histogram of causal allele frequencies in EUR and YRI. (B) CDF of causal allele frequencies in EUR and YRI.**

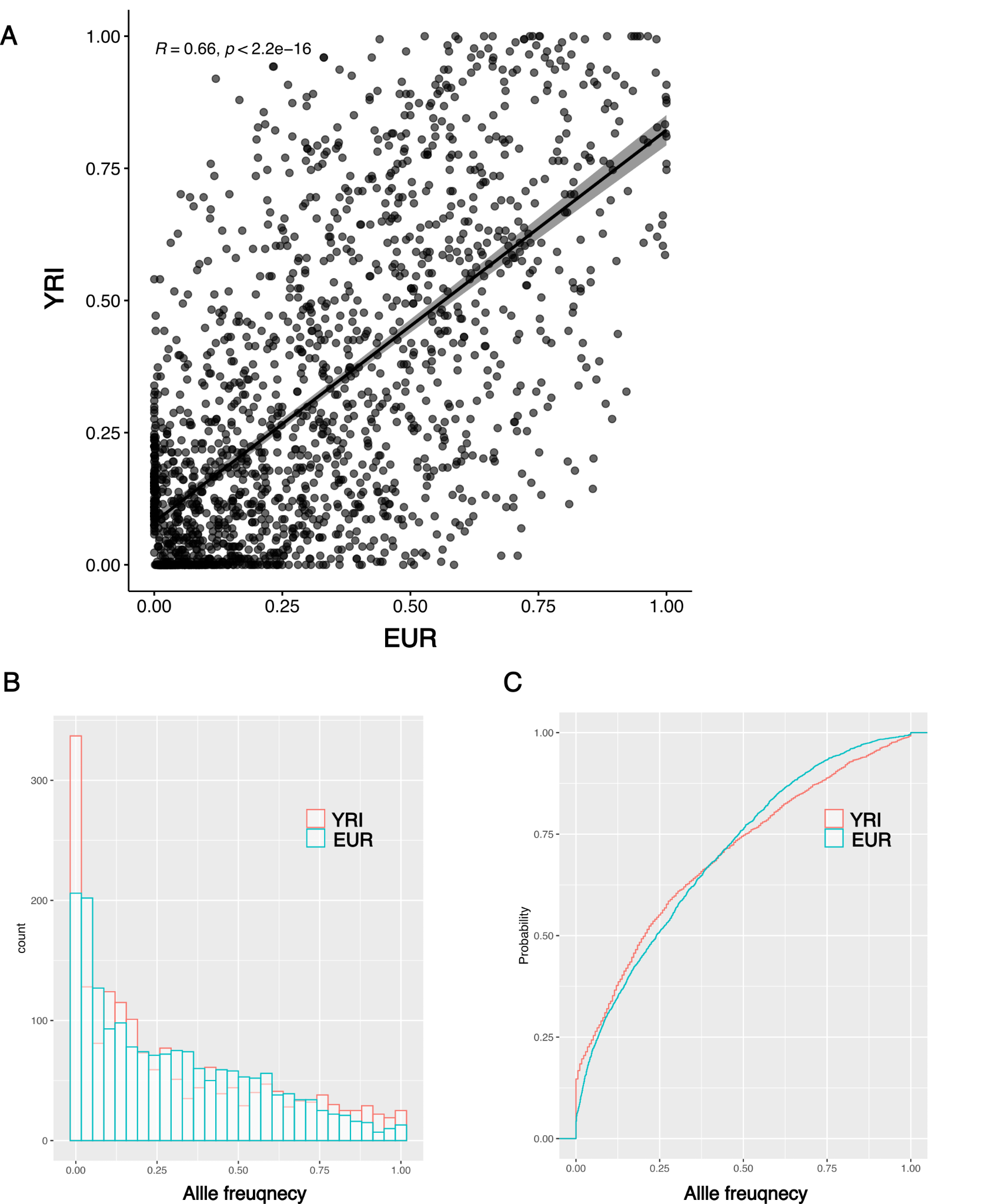

Supplementary Tables (all supplementary tables are in Additional File 1)

**Table S1** Genetic correlation, heritability, h2_hom and h2_het of genes

**Table S2** R2 and portability of genetic scores of all investigated genes

**Table S3** Max varLD scores of all investigated genes

**Table S4** Differential gene expression in EUR and YRI

**Table S5** Frequency of the expression-increasing allele of the fine-mapped variants in EUR and YRI
